# Examining the complex dynamics influencing acute malnutrition in Turkana and Samburu counties: study protocol

**DOI:** 10.1101/2024.08.18.24312202

**Authors:** Calistus Wilunda, Estelle Sidze, Faith Thuita, Dickson Amugsi, Amanuel Abajobir, Martin Mutua, Bonventure Mwangi, Samuel Iddi, Chessa Lutter, Valerie Flax, Albert Webale, Esther Anono, Hazel Odhiambo, Caroline Wangui Wainaina, Elizabeth Kimani-Murage, Stephen Ekiru, Gillian Chepkwony, John Ebei, Duncan Lesiamito, Brad Sagara

**Author notes:** Corresponding author: African Population and Health Research Center, P.O. Bo× 10787-00100, APHRC Campus, Kitisuru, Nairobi, Kenya.

## Abstract

Acute malnutrition in children under 5 years is persistent in Eastern Africa’s arid and semi-arid lands. This study aimed to identify the drivers of acute malnutrition in Turkana and Samburu counties, Kenya. This was a population-based longitudinal mixed-methods observational study. Qualitative and quantitative data were collected at Wave 1 but only quantitative data were collected during follow-up. Participants were a representative sample of children and their primary caregivers from households with children under 3 years at Wave 1. Anthropometric measurements of all children under 5 years in the sampled households were taken at Wave 1 (May to July 2021), and one child under 3 years was randomly selected for follow-up about every 4 months over 2 years for six data collection waves. The study also collected data on socio-demographics; child feeding practices and morbidity, household water and food insecurity; shocks; coping strategies, social safety nets and economic safeguards; water, sanitation, and hygiene; women’s decision making; and food consumption. Qualitative data were collected through community dialogues, focus group discussions, in-depth interviews, photovoice and key informant interviews with mothers and fathers with children under 5 years, community leaders, county officials and staff of nongovernmental organizations. The data were coded and analyzed thematically. Data collection is complete, and analysis is ongoing. The analysis includes thematic analysis of qualitative data and descriptive and multivariable regression analysis of quantitative data.

## Introduction

Acute malnutrition in infants and children under 5 years of age is persistent in the arid and semi-arid lands (ASALs) of Eastern Africa despite years of investment by the United States Agency for International Development (USAID), other donors and the government. In the ASALs of Kenya, including Samburu and Turkana counties, the situation is exacerbated by deep-rooted poverty, socio-cultural practices, unequal access to basic services, migration, and poor seasonal rainfall/droughts and other shocks. Children are the most affected by poor nutrition mainly because of systemic/basic, underlying, and immediate factors as illustrated in the conceptual framework for acute malnutrition in Africa’s drylands (H. Young, 2020). Systemic/basic factors include inadequate empowerment of women and limited control over household resources; high workload; domestic violence; and alcohol consumption. Underlying factors include household food insecurity; inadequate social and care environment; inadequate health services; and an unhealthy environment. Immediate factors include inadequate dietary intake and high disease burden (Shell-Duncan & Obungu Obiero, 2000; H. Young, 2020).

Children living in extreme poverty, as is the case in many parts of Turkana and Samburu, face increased risks of undernutrition, poor health, and suboptimal cognitive development (Worku et al., 2018). Poverty and low parental education are established risk factors for child undernutrition (Adeyeye, Adebayo-Oyetoro, & Tiamiyu, 2017; Vollmer, Bommer, Krishna, Harttgen, & Subramanian, 2016). Standardized Monitoring and Assessment of Relief and Transitions (SMART) surveys from Turkana and Samburu show very high levels of illiteracy. For example, in 2019, 78.8% and 68.8% of household heads in Turkana and Samburu, respectively, had no formal education (Samburu County Department of Health., 2019; Turkana County Department of Health., 2019). The surveys also showed high prevalence of preventable and treatable childhood illnesses, such as acute respiratory infection, diarrhea, and malaria/fever (Samburu County Department of Health., 2019; Turkana County Department of Health., 2019), which increase the risk of undernutrition. The prevalence of global acute malnutrition [weight for weight-for-height z-scores (WHZ) <-2 SD] among children under 5 years in the two counties is consistently at or above the emergency threshold of 15% (Samburu County Government, 2019; Turkana County Government, 2020), with no improvement over the past three decades (KNBS & ICF, 2023).

Virtually no evidence existed on how nutrition outcomes and related variables change over time and seasons within the same households and the synergistic effects of frequent and severe climate-related shocks. Tracking acute malnutrition and its associated factors longitudinally was designed to assess which of these factors are associated with changes in acute malnutrition over two years, and the relative strength of those associations. An initial desk review on the status and key drivers of acute malnutrition informed the decision to conduct a longitudinal study with two main objectives: 1) to understand how a variety of immediate, underlying, and systemic/basic factors influence acute malnutrition across seasons among young children living in four different livelihood zones; and 2) to identify and prioritize opportunities and barriers to achieve sustained reductions in acute malnutrition.

Nawiri is a USAID-funded program aimed at reducing persistent acute malnutrition among children in Turkana and Samburu counties. Mercy Corps leads the consortium consisting of Research Triangle International (RTI), African Population and Health Research Center (APHRC), Save the Children, BOMA, and Caritas to implement the project. These consortium partners handle different components of the project, with APHRC and RTI leading the longitudinal study. The present study was conceptualized in consultation with the government and other key stakeholders in Turkana and Samburu counties.

### Key Messages

- Acute malnutrition in children under 5 years is persistent in Eastern Africa’s arid and semi-arid lands.
- Nawiri Longitudinal study, which aimed to identify the drivers of acute malnutrition in Turkana and Samburu counties, Kenya, is a population-based longitudinal mixed-methods observational study.
- Participants were a representative sample of children and their primary caregivers from households with children under 3 years at Wave 1.
- Index children were followed about every 4 months over 2 years for six data collection waves.
- Study findings are informing program design, implementation, and adaptation to prevent acute malnutrition and improve the counties’ nutrition surveillance.

## Methods

### Settings

This study was conducted in the neighboring counties of Turkana and Samburu in northern Kenya (**Figure 1**). Turkana County is the second largest of Kenya’s 47 counties with a total area of about 71,600 km^2^ and is divided into seven sub-counties: Turkana Central, Loima, Turkana South, Turkana East, Turkana North, Kibish, and Turkana West. The county had 926,976 residents according to the 2019 national census (Kenya National Bureau of Statistics, 2019a). Samburu County has a land area of approximately 21,022 km^2^ and is divided into three sub-counties: Samburu Central, Samburu North, and Samburu East. The county’s population was 310,327 according to the 2019 national census (Kenya National Bureau of Statistics, 2019a). Both counties have a hot and dry climate, unreliable rainfall patterns, frequent droughts and are classified as ASALs, with 80% of Turkana being categorized as either arid or very arid. Approximately 60% of the population in Turkana is pastoral, 20% is agropastoral, 12% are fisher folks and 8% is engaged in formal or informal employment in urban/peri-urban areas (Turkana County Government, 2019). Turkana is the poorest county in Kenya, with 77.7% of the population living below the poverty line in 2021 compared to a national average of 38.6% (Kenya National Bureau of Statistics, 2023). The main livelihoods in Samburu are pastoralism (57%), agropastoralism (37%) and petty trade (6%)(Samburu County Government, 2019). **Table 1** summarizes the characteristics of the two counties.

**Figure 1:**
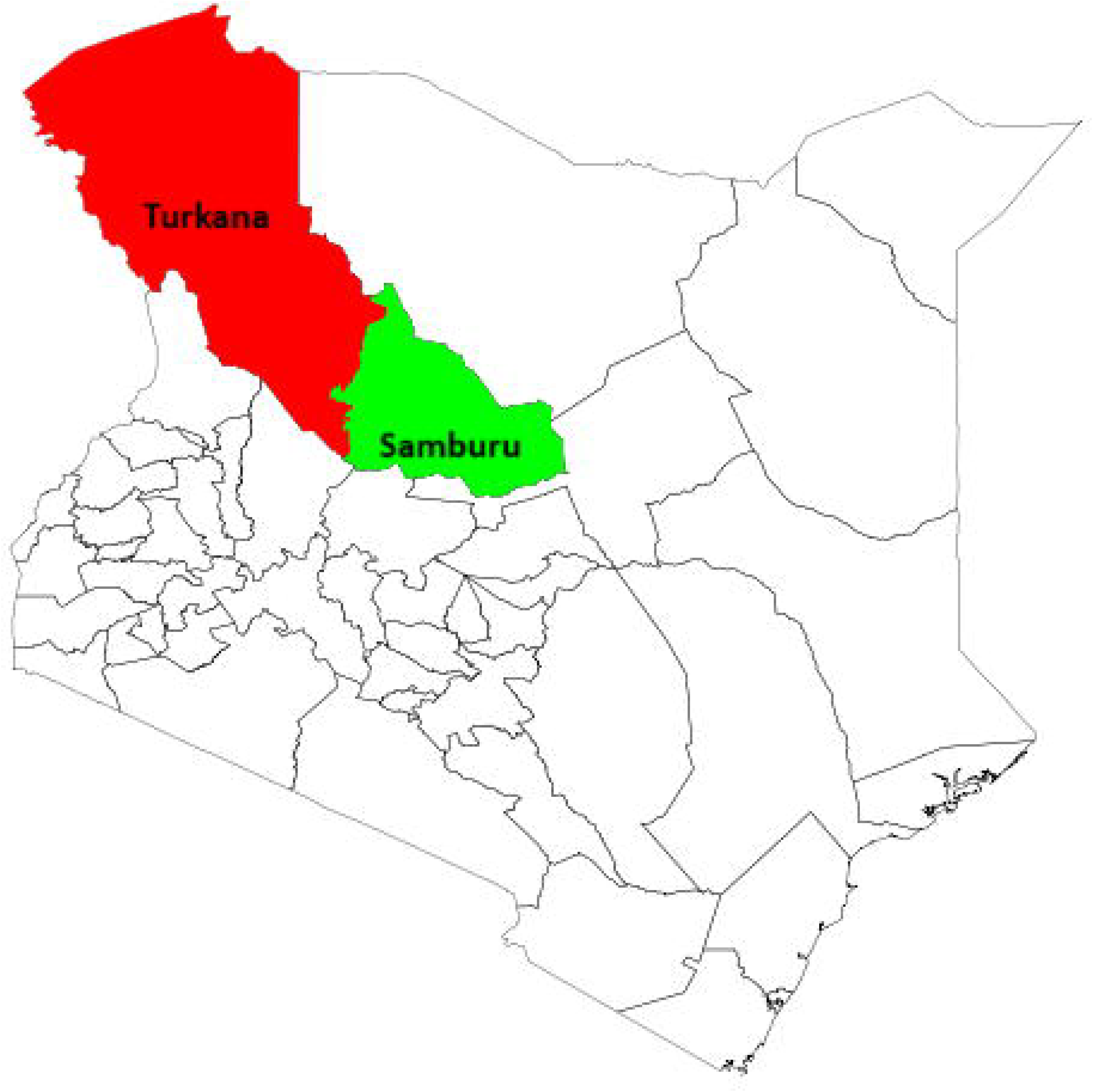
Map of study counties

**Table 1.**
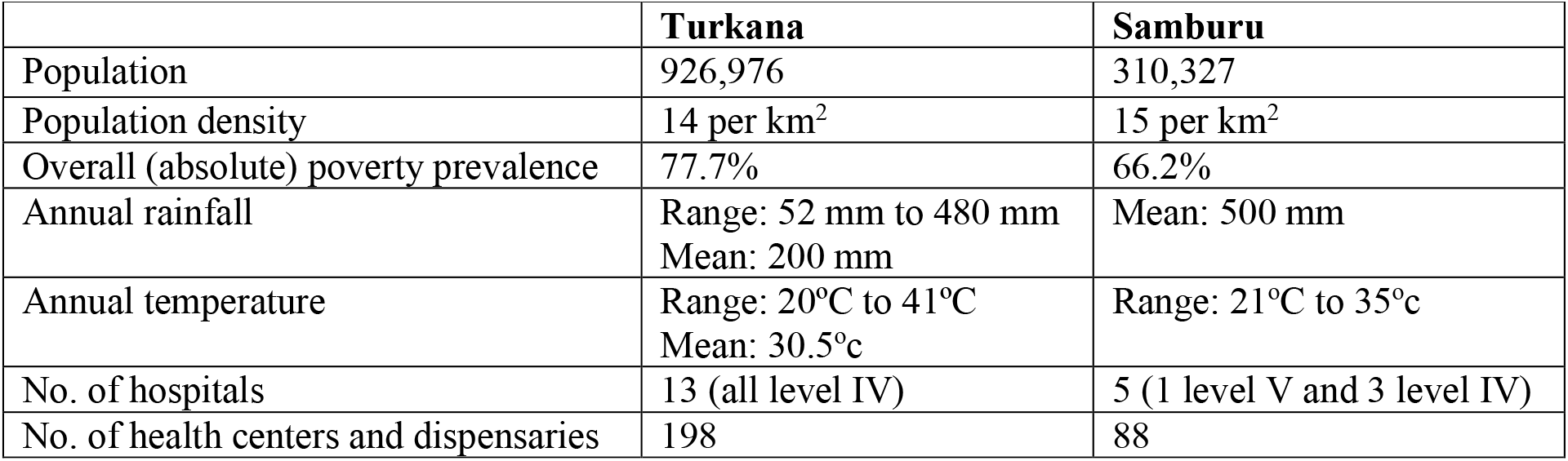
A summary of the characteristics of the study counties.

|  | **Turkana** | **Samburu** |
| --- | --- | --- |
| Population | 926,976 | 310,327 |
| Population density | 14 per km<sup>2</sup> | 15 per km<sup>2</sup> |
| Overall (absolute) poverty prevalence | 77.7% | 66.2% |
| Annual rainfall | Range: 52 mm to 480 mm<br>Mean: 200 mm | Mean: 500 mm |
| Annual temperature | Range: 20°C to 41°C<br>Mean: 30.5°C | Range: 21°C to 35°C |
| No. of hospitals | 13 (all level IV) | 5 (1 level V and 3 level IV) |
| No. of health centers and dispensaries | 198 | 88 |

### Study Design and Participants

This was a 24-month longitudinal mixed-methods (quantitative and qualitative) observational study of children under 3 years at recruitment and their mothers or primary caregivers (where the mother was not available). Eligible children and their mothers/caregivers were recruited and followed every 4 months for six waves of data collection. Anthropometric measurements (weight, length/height, and MUAC) of all children under 5 years in the sampled households were taken at baseline (Wave 1), and one child under 3 years per household (the index child) was selected and followed up through Wave 6. In case there was more than one child under 3 years in the household, the index child was selected randomly.

The qualitative component of the study was implemented at Wave 1 and included community dialogues, key informant interviews (KIIs), focus group discussions (FGDs), in-depth interviews (IDIs), and photovoice – a community-based participatory and visual research method where participants use photography to capture and express the issues affecting them within their environment (Wang, 1999). Community dialogues were conducted separately for men and women of different age groups to mitigate power dynamics and maximize self-expression. The women’s groups included adolescent mothers (10–17 years), younger mothers (18–24 years), and older mothers (25+ years). The men’s groups included younger fathers (15–24 years) and older fathers (25+ years). All community dialogue participants had a child aged 3 years or younger. Each community dialogue had 24 participants. The community dialogue included three methods: (1) a free listing of factors that the participants viewed as contributing to acute malnutrition; (2) a seasonal calendar, used to explore and understand how different seasonal factors influence malnutrition from the communities’ perspective; and (3) causal mapping, used to map community perceptions of the connections between factors said to contribute to acute malnutrition. The free listing was done by all participants together and thereafter the participants were divided into four groups by age and gender (i.e., adolescent and younger mothers, older mothers, younger fathers, and older fathers) to perform the seasonal calendar and causal mapping activities.

KIIs were conducted with county government officials, community gatekeepers, representatives of local non-governmental organizations (NGOs), United Nations agencies, USAID Partnership for Resilience and Economic Growth partners, and national government officials. These respondents offered expert insights into the nutritional situation and related community and environmental factors influencing acute malnutrition and how it can be addressed.

FGDs and IDIs were conducted with adolescent mothers, younger mothers, older mothers, younger fathers, and older fathers, all with children aged 3 years or younger. Separate IDIs were conducted with adolescent mothers, younger mothers, and older mothers with acutely malnourished children aged 2 years or younger. The FGDs and community dialogue discussions were undertaken separately for both men and women. The FGDs and IDIs were used to collect information on community perceptions of factors related to acute malnutrition; maternal, infant, and young child nutrition (MIYCN) practices; water, sanitation, and hygiene practices; and health-seeking behaviors. Additionally, FGD respondents participated in a free listing activity to collect information on what constitutes an enabling environment for mothers to achieve optimal MIYCN practices, what support networks already exist to promote optimal MIYCN practices for community women, and how the enabling environment and networks could be enhanced.

Photovoice exercise aimed to understand behaviors that drive acute malnutrition, and included adolescent mothers (10-17 years), younger mothers (18-24 years), and older mothers (25+ years).

### Sample Size and Sample Allocation

The sample size for the quantitative component was estimated separately for Turkana and Samburu using the household survey sample size formula by the United Nations Statistical Division (United Nations., 2008).

For Turkana, we assumed an under-3 years global acute malnutrition (GAM) prevalence of 23.2%, design effects of 1.5 due to stratification and clustering (Turkana County Department of Health., 2019) and 1.12 to account for repeated measurements on the same individuals at six time points, a common correction of 0.02 based on a previous study that estimated an intraclass correlation of 0.0044 for clustering of children within a household (Becquey & et al., 2019), a margin of error of ± 5 percentage points, 95% confidence interval (CI), a nonresponse and attrition rate of 20%, the proportion of the population targeted for the study (children under 3 years) at 7.6% and average household size of six (Kenya National Bureau of Statistics, 2019a, 2019b). The minimum required sample size was 1,544 households.

For Samburu, we assumed an under-3 years GAM prevalence of 12.5%(Samburu County Department of Health., 2019), design effects of 1.3 due to stratification and clustering (Samburu County Department of Health., 2019) and 1.12 to account for the repeated measurements on the same individuals at six time points, a common correction of 0.02 based on a previous study, a margin of error of ± 5 percentage points, 95% CI, a nonresponse and attrition rate of 20%, the proportion of the population targeted for the study at 9.2%, and the average household size of five (Kenya National Bureau of Statistics, 2019a). The minimum required sample size was 669 households.

The number of households was allocated proportionally to the population size of each stratum (or survey zone), as shown in **Table 2**. In each stratum, a random sample of 25 and 20 villages in Turkana and Samburu, respectively, was selected using probability proportional to the population size of the cluster. Equal numbers of households were then sampled from each village in each of the survey zones (Turkana: Central 20, North 7, West 16, and South 20; and Samburu: North 8, Central 18, and East 9) to ensure that each of the households had the same probability of being selected. In some villages, the target number of households was not achieved because of insecurity or immigration and a spare village was sampled randomly from the remaining villages to achieve the target sample size. All the 2,267 and 664 villages in Turkana and Samburu counties, respectively, were considered.

**Table 2.** Sample size allocation for the quantitative component.

| County and survey zone | Administrative sub-counties | Population | Villages | Proportional allocation | Sampled villages |
| --- | --- | --- | --- | --- | --- |
| Turkana |  |  |  |  |  |
| Turkana Central | Central and Loima | 293,100 | 902 | 488 | 25 |
| Turkana North | North and Kibish | 101,987 | 379 | 170 | 25 |
| Turkana West | West | 239,627 | 425 | 399 | 25 |
| Turkana South | South and East | 292,262 | 561 | 487 | 25 |
| Total |  | 926,976 | 2267 | 1544 | 100 |
| Samburu |  |  |  |  |  |
| Samburu West | Samburu Central | 164,942 | 366 | 356 | 20 |
| Samburu East | Samburu East | 67,391 | 111 | 160 | 20 |
| Samburu North | Samburu North | 77,994 | 187 | 179 | 20 |
| Total |  | 310,327 | 664 | 695 | 60 |

A total of 96 KIIs, 14 community dialogues with 319 participants, 35 FGDs with 211 participants, 138 IDIs, and 21 photovoice sessions with 126 participants were conducted across all livelihood zones in Turkana and Samburu. **Table 3** summarizes the number of participants and sample allocation for each qualitative data collection activity by county and livelihood zone.

**Table 3.** Sample size and sample allocation for the qualitative component by county and livelihood zone.

|  | Turkana |  |  |  |  | Samburu |  |  |  |
| --- | --- | --- | --- | --- | --- | --- | --- | --- | --- |
|  | Pastoral | Agro-pastoral | Fisher folk | Urban/peri-urban | Total | Pastoral | Agro-pastoral | Urban/peri-urban | Total |
| **Key informant interviews** |  |  |  |  |  |  |  |  |  |
| Community leaders | 9 | 9 | 8 | 9 | 35 | 8 | 8 | 13 | 29 |
| County officials and NGO representatives | - | - | - | - | 14 | - | - | - | 18 |
| **Community dialogues** |  |  |  |  |  |  |  |  |  |
| Mothers (all ages) | 1 | 1 | 1 | 1 | 4 | 1 | 1 | 1 | 3 |
| Fathers (all ages) | 1 | 1 | 1 | 1 | 4 | 1 | 1 | 1 | 3 |
| **Total number of participants** | **42** | **48** | **42** | **48** | **180** | **48** | **47** | **44** | **139** |
| **Focus group discussions** |  |  |  |  |  |  |  |  |  |
| Adolescent mothers (10–17 years) | 1 | 1 | 1 | 1 | 4 | 1 | 1 | 1 | 3 |
| Younger mothers (18–24 years) | 1 | 1 | 1 | 1 | 4 | 1 | 1 | 1 | 3 |
| Older mothers (25+ years) | 1 | 1 | 1 | 1 | 4 | 1 | 1 | 1 | 3 |
| Younger fathers (15–24 years) | 1 | 1 | 1 | 1 | 4 | 1 | 1 | 1 | 3 |
| Older fathers (25+ years) | 1 | 1 | 1 | 1 | 4 | 1 | 1 | 1 | 3 |
| Total number of groups | 5 | 5 | 5 | 5 | 20 | 5 | 5 | 5 | 15 |
| **Total number of participants** | **31** | **30** | **30** | **30** | **121** | **30** | **30** | **30** | **90** |
| **In-depth interviews** |  |  |  |  |  |  |  |  |  |
| Adolescent mothers (10–17 years) | 3 | 5 | 4 | 4 | 15 | 3 | 6 | 5 | 14 |
| Younger mothers (18–24 years) | 4 | 6 | 6 | 8 | 18 | 4 | 2 | 4 | 10 |
| Older mothers (25+ years) | 6 | 6 | 7 | 8 | 26 | 7 | 7 | 5 | 19 |
| Younger fathers (15–24 years) | 2 | 2 | 2 | 2 | 8 | 2 | 2 | 2 | 6 |
| Older fathers (25+ years) | 2 | 2 | 2 | 2 | 8 | 2 | 2 | 2 | 6 |
| **Total number of participants** | **17** | **21** | **21** | **24** | **83** | **18** | **19** | **18** | **55** |
| **Photovoice** |  |  |  |  |  |  |  |  |  |
| Adolescent mothers (10-17 years) | 1 | 1 | 1 | 1 | 4 | 1 | 1 | 1 | 3 |
| Younger mothers (18-24 years) | 1 | 1 | 1 | 1 | 4 | 1 | 1 | 1 | 3 |
| Older mothers (25+ years) | 1 | 1 | 1 | 1 | 4 | 1 | 1 | 1 | 3 |
| Total number of groups | 3 | 3 | 3 | 3 | 12 | 3 | 3 | 3 | 9 |
| **Total number of participants** | **18** | **18** | **18** | **18** | **72** | **18** | **18** | **18** | **54** |

### Sampling

A representative sample of children under 3 years and their mothers/caregivers was obtained using a multistage sampling approach, with survey zones as units of stratification. In Turkana, Nawiri designated four survey zones (Central, North, West, and South) that include all the livelihood zones (pastoral, agro-pastoral, fisher folk, and urban/peri-urban). In Samburu, there are three survey zones (North, Central, and East) covering all three livelihood zones (pastoral, agro-pastoral, and urban/peri-urban). The livelihood zones were delineated to generate evidence on the unique vulnerabilities of communities pursuing different livelihood strategies. Villages were treated as clusters within a survey zone, from which a random sample of 25 and 20 villages in Turkana and Samburu, respectively, was drawn. A household listing was conducted in each of the selected villages to identify and enumerate all households with children under 3 years old. Households with at least one child under 3 years old formed the sampling frame for the final stage of random selection of households. For sampled households with more than one child under 3 years, one child was selected randomly to participate in the study.

For the qualitative component, four villages were purposefully selected in consultation with county officials to represent the different livelihood zones (one village per livelihood zone) and sub-counties **(Table 4)**. In each village, participants were recruited with the help of the chief/assistant chief and the community health volunteer and invited to provide informed consent and participate in data collection.

**Table 4.** Villages selected for qualitative study, by county and livelihood zone.

| County and sub-county | Location | Sub-location | Village | Livelihood zone |
| --- | --- | --- | --- | --- |
| **Turkana** |  |  |  |  |
| Central | Kangathotha | Namukuse | Nakwamekwi | Fisherfolk |
| Central | Township | Nakwamekwi | Nanyangakipi | Urban/peri-urban |
| South | Lokichar | Naposmoru | Kekorongor | Pastoral |
| East | Katilia | Katilia | Kanakipe B | Agro-pastoral |
| **Samburu** |  |  |  |  |
| Central | Loosuk | Loosuk | Loisukutan | Agro-pastoral |
| Central | Central | Maralal | Maralal | Urban/peri-urban |
| North | Ndoto | Ndoto | Tankar A | Pastoral |

### Data Collection and Data Management

#### Quantitative

Wave 1 data were collected in May–June 2021, during long rains in Turkana and June – July 2021, during a continental rainy season in Samburu. Supplementary **Figure S1** shows the timing of data collection and seasons for all waves. There was a prolonged drought at Waves 3-5 in Turkana and at Waves 3 and 4 in Samburu.

Data were collected by experienced and trained fieldworkers using a questionnaire loaded in SurveyCTO – a survey platform for electronic data capture that has in-built skips and quality checks to increase efficiency and reduce the time needed for data cleaning. The questionnaire included the following modules: (1) identification and tracking, (2) demographics and household composition, (3) anthropometry of children under 5 years and mothers, (4) socioeconomics, (5) household food security, (6) water, sanitation and hygiene (WASH), (7) morbidity and health-seeking behaviors, (8) MIYCN, (9) shock experience/exposure, (10) shock preparedness and response, and (11) caregiver’s psychological well-being. Supplementary **Table S1** presents a description of the indicators, methods, and frequency of data collection for survey data at each wave. Data were uploaded from the tablets onto a secure APHRC server after each day of data collection. Data were synchronized automatically to a server when the tablet was in a location with internet network coverage. The uploaded data were then checked daily for quality by a data manager and a team dedicated to coordinating field procedures in both counties and at the APHRC head office in Nairobi.

At baseline, fieldworkers were recruited and trained in data collection processes for 10 days. A three-day refresher training was conducted at every subsequent wave of data collection. All fieldworkers were recruited from the local communities because they were familiar with the area, customs, and local languages. They were trained in the overall aim of the study, study tools, research ethics, interviewing techniques, and the process for obtaining informed consent. Mock interviews, field-based pilots, and debrief sessions after the pilots were conducted. The fieldworkers were also trained in the use of tablet-based questionnaires and anthropometric measurement techniques.

In addition to in-built quality control measures in the tablet-based platform and fieldworker training, other data quality control measures included conducting regular spot checks and sit-ins, mirror interviews and observation of interviews and anthropometric data measurement procedures by supervisors, data review by the field coordinators, regular data quality checks by the data manager, supportive supervision by county officials, and post-survey data cleaning. Field data collection processes in each wave were also monitored by independent teams comprising of government technical officers and Nawiri monitoring and evaluation staff.

Anthropometric equipment were calibrated daily during fieldwork to ensure accurate measurements. The weight of children was measured using a calibrated digital electronic mother/caregiver–child pair weighing scale (Seca 874–200kg). For very young children or those who could not stand on the weighing scale, weight was measured using tared weighing, whereby the weight of the mother or caregiver was measured first, after which she was asked to hold the child and stand on the scale. The mother’s or caregiver’s weight was then subtracted from the combined weight of the mother/caregiver and child to obtain the child’s weight. Adult and child height were measured using a stadiometer (Seca 213–220 cm) and length board (wooden: length/height to 130 cm), respectively. The measuring boards were calibrated using piping of a known length, while each scale was tested with a standard weight of 5 kg. Child and adult MUAC were measured to the nearest 0.1 cm using United Nations Children’s Fund (UNICEF)-simplified MUAC tapes that showed three classes: red, yellow, and green. Each of the weight and height measurements was taken twice and an average was calculated to ensure accuracy. Respondents were asked to remove all excess clothing and other items. Data on edema were collected for all children. Fieldworkers were trained on how to identify children with the condition, using visual aids.

The primary outcome variable was GAM (weight-for-height z-score [WHZ] < –2 standard deviations [SD], MUAC < 125 millimeters [mm]) or MUAC-for-age z-score (< –2 SD). The secondary outcome variables were stunting (height-for-age z-score [HAZ] < –2 SD) and underweight (weight-for-age z-score [WAZ] < –2 SD) (WHO Multicentre Growth Reference Study Group, 2006).

The height and weight of mothers and caregivers were used to compute body mass index (BMI). The BMI was computed by dividing weight (in kilograms [kg]) by height in meters squared (m2) and categorized into underweight (BMI < 18.5 kg/m^2^), normal weight (BMI = 18.5–24.99 kg/m^2^), overweight (BMI = 25–29.99 kg/m^2^), and obese (BMI ≥30 kg/m^2^).

Underweight among pregnant women was assessed using MUAC with a cut-off value of 21 centimeters (cm), and short stature was assessed using the cut-off value of 145 cm as recommended by the Pan American Health Organization (Pan American Health Organization, 1991).

Infant and young child feeding practices were assessed using indicators from the World Health Organization and UNICEF (WHO & UNICEF, 2021). Minimum dietary diversity for women was determined using a cut-off value of 5 out of the 10 food groups recommended by the Women’s Dietary Diversity Project Study Group (Women’s Dietary Diversity Project Study Group, 2017).

A reduced coping strategy index (rCSI) was calculated using a set of behaviors with a universal set of severity weightings for each behavior (Maxwell & Caldwell, 2008). The five standard coping strategies and their severity weightings used in rCSI calculation included eating less-preferred foods (1.0), borrowing food or money from friends and relatives (2.0), limiting portions at mealtime (1.0), limiting adult food intake (3.0), and reducing the number of meals per day (1.0).

At baseline, a household wealth index was created using principal component analysis based on ownership of assets; house wall, floor, and roof materials; and light source. The index was then used to rank households into wealth tertiles or quintiles. Also, information on household size, sex of household head, education of household head, household adult composition (male and female adults, female adults only, male adults only or no adults), and household wealth was collected at Waves 1 and updated at 6.

Household poverty likelihood (the probability that the household is poor) was calculated based on household responses to 10 poverty probability index (PPI) questions (Innovations for Poverty Action, 2018). A PPI score was obtained by adding up the points allotted to the responses given by the household. This was then converted to a poverty likelihood by referring to published tables for the 2015 Kenya PPI (Innovations for Poverty Action, 2018).

Household food insecurity was measured using the household Food Insecurity Experience Scale (FIES) (Ballard, Kepple, & Cafiero, 2013), which consists of eight questions capturing a range of food insecurity severity, with yes/no responses. FIES data were analyzed by Rasch modelling (Nord, 2014) based on the principles of item response theory using the RM.weights package in R (Cafiero, Viviani, & Nord, 2028) to obtain food insecurity prevalence rates.

Household water insecurity was measured using the Household Water Insecurity Experiences (HWISE) Scale, which consists of 12 items each with four response categories (S. L. Young et al., 2019). The total score, which is obtained by summing up scores of the 12 questions, can range from 0 to 36, and a household with a score of ≥12 is classified as water insecure. Data were also collected on other WASH indicators including toilet facility, hand washing, distance to water source, type of water source, and water treatment.

#### Qualitative

Data were collected face-to-face with eligible participants, using qualitative guides for each method of data collection (i.e., FGD, KII, IDI, or community dialogue) and type of participant by teams of fieldworkers from May 10 to June 30, 2021. All fieldworkers were recruited from their communities and were familiar with the local culture and spoke the local languages. They were trained intensively using APHRC’s training protocol, which included both theory and practice. The training also covered the use of audio recording devices and taking field notes to capture nonverbal cues, observations, and other relevant information. The fieldworkers were initially trained for 7 days in March 2021; however, due to delays in commencing fieldwork, a 2-day refresher training was offered to all fieldworkers in May 2021 before the data collection. A pilot was carried out immediately after the training to test the tools and field procedures.

Supervision consisted of two layers: (1) daily supervision by team leaders to ensure that the recruited participants met the target criteria, fieldworkers conducted the assigned interviews, recorded data were backed up daily, and debriefing meetings were conducted; and (2) weekly reviews by the data coordination team to ensure that all interviews were conducted.

Data quality control measures included random observation of group discussions or interviews, daily debriefing meetings by team leaders, cross-checking field summary notes, and holding weekly debriefing meetings between the APHRC research team and fieldworkers to review the quality of the incoming data. Audio recordings were backed up daily on a secure computer. The APHRC research team cross-checked 10% of transcribed audio files against the guide to ensure that all research questions were answered.

The photovoice exercise was carried out in selected villages across all livelihoods. Each photovoice session had six members who were split into two groups to take photos, but discussions were undertaken by the whole group stratified by age. Photovoice activities involved training research assistants who in turn trained participants on the ethics and use of cameras. This was followed by provision of cameras to selected participants, who were asked to spend 1-2 days photographing their lived experiences and perspectives on factors influencing acute malnutrition in their communities, including food, MIYCN, WASH, and women’s activities while exploring possible solutions. Under the moderation of research assistants, participants then reviewed the photographs, selected the relevant ones for printing and provided captions and descriptions explaining what each of the photographs represented.

All audio files were cross-checked for standard labeling and to confirm the achievement of the set targets per livelihood zone and group. The audio files (for IDIs, FGDs, KIIs, and community dialogues) were then transcribed verbatim from local languages and translated into English by a team of experienced transcribers with good mastery of local languages and English. Content analysis and thematic groupings were undertaken by experienced independent coders. After reading and rereading the transcripts, the coders developed a codebook of deductive and inductive codes for each interview guide.

### Data analysis

#### Quantitative data

A detailed data analysis plan was developed before data collection. Because this study was designed to generate evidence to inform the development and adaptation of Nawiri programmatic interventions and strategies to reduce acute malnutrition, preliminary data analysis was conducted at the end of each wave. Descriptive analysis of the characteristics of children; mothers/caregivers; and households,WASH and MIYCN indicators, household food insecurity and coping strategies, morbidity and health-seeking behaviors, shock experience/exposure and economic safeguards, and women’s decision making was conducted at baseline and each subsequent wave of data collection. A preliminary analysis of the determinants of acute malnutrition was performed at Waves 1 and 3 and a final analysis after completion of Wave 6 is ongoing. The analysis, which is performed in Stata and accounts for the complex survey design, includes both bivariate and multivariable analysis using mixed-effects regression models to handle repeated measurements and time-invariant and time-varying predictors. Mixed effects models also handle irregularly timed and missing data without the need for explicit imputation.

#### Qualitative

Transcripts from IDIs, FGDs, KIIs, and community dialogues were cleaned, saved in rich-text format for importation into NVivo (QSR International) software, and analyzed according to the developed codebook using a thematic framework approach (Pope, Ziebland, & Mays, 2000). Intercoder variability in thematic groups was limited to less than 10%. Transcripts from the seasonal calendar and causal mapping exercises were coded and analyzed thematically. Rankings of factors in the seasonal calendars were analyzed as counts and compiled into graphs by livelihood zone. Information collected through the causal mapping exercise was presented visually using a causal diagram to demonstrate community perceptions of linkages between factors, their relative importance, and the livelihood zones that mentioned them.

The free listing data from FGDs were analyzed by grouping responses from participants on the different items of support for optimal MIYCN and then tabulating the frequencies. This step was followed by consolidating the responses further into broader thematic areas. The detailed description of the broader themes was derived from the transcripts of the discussions during the free listing exercises.

## Discussion

This paper describes the design and implementation of a 24-month longitudinal mixed-methods observational study of young children and their mothers or primary caregivers in Turkana and Samburu counties in Kenya. The study was designed to generate contextual and nuanced information to support the USAID Nawiri program strategy design and co-creation of integrated interventions to sustainably reduce acute malnutrition. The study adopted a systems-based approach to surface contextualized evidence to inform the design and iterative adaptation of a multi-sector program for sustainably reducing acute malnutrition in the counties. Data collection is complete, and analysis is ongoing. Challenges encountered in the implementation of this study included inadequate mobilization of respondents in some villages leading to initial delays in the household listing, respondents’ fatigue, out-migration and insecurity in some areas, long distances between households and tough terrain and impassable roads due to heavy rains in some areas forcing fieldworkers to walk long distances to access the villages, absence of respondents, and the COVID-19 pandemic, which led to significant delays in data collection.

## Supporting information

Supplementary Table and Fugure

## Data Availability

This is a study protocol and has no underlying data.

## Ethical statement

Ethical and research approvals and research permits were obtained from Amref Health Africa’s Ethical and Scientific Review Committee (Amref ESRC P905/2020) and the National Commission for Science, Technology, and Innovation of Kenya, respectively. A reliance agreement was signed between the institutional review boards at APHRC and RTI. Informed consent was obtained from all participants during Wave 1 and reconfirmed at Waves 2 to 6. To ensure confidentiality in the qualitative component, the research team saved transcripts and photos on a dedicated password-protected computer and removed all identifiers from the transcripts. Only anonymized transcripts will be kept in the APHRC data repository. Informed photo-release consent was obtained to allow the use of photos in reports, publications, websites, presentations, exhibitions, and any other future use related to the study. During data collection, all COVID-19 risk-reduction guidelines issued by the Government of Kenya were followed. Also, all survey staff and participants wore face masks, and hand sanitizer was made readily available.

## Author contributions

ES, FT, DA, AA, MM, CL, VF, AW, and BS designed the study. CW, DA, AA, MM, BM, EA, HO, CWW, SE, GC, JE, and DL implemented fieldwork. CW, ES, FT, and EK-M provided study oversight. CW prepared the first draft of the manuscript and incorporated comments from the co-authors. All authors contributed to the revision of the manuscript and approved the final version.

