## Supplementary Table and Fugure for "Examining the complex dynamics influencing acute malnutrition in Turkana and Samburu counties: study protocol"

**Supplementary files**

**Table S1:** Indicators, methods, and frequency of data collection for the quantitative component.

| Variable/indicator | Method | Frequency of data collection |
| --- | --- | --- |
| Identification and tracking   - Global positioning system (GPS) coordinates - Village name - Names and contact information of primary adults in the household | Survey | Wave 1 with checks for any changes in subsequent waves |
| Demographics and household composition   - Number of household members, number of children under 5 years, maternal/paternal education, paternal/maternal occupation, ethnicity, religion, etc. | Survey | Wave 1 with checks for any changes in subsequent waves |
| Anthropometry of children under 5 years and mothers or caregivers   - Mid-upper arm circumference (MUAC) - Weight - Length/height | Survey | At all survey waves |
| Socioeconomics   - Household wealth - Livelihoods (household asset base, income sources, social protection, livestock number, access to markets, access to land/pasture) - Household decision-making and control over resources - Poverty probability index | Survey | At all survey waves, with some modifications for indicators unlikely to vary sub-annually |
| Household food security   - Coping Strategies Index (CSI), - Household Food Insecurity Experience Scale (HFIES) | Survey | At all survey waves |
| Water, sanitation, and hygiene (WASH)   - Water source, access, availability, and seasonality - Household Water InSecurity Experiences (HWISE) scale - Hygiene practices | Survey | At all survey waves |
| Health-seeking behavior   - Integrated Management of Acute Malnutrition (IMAM) - Community health service experience - Child morbidity | Survey | At all survey waves, with some modifications for indicators unlikely to vary sub-annually |
| Maternal, infant, and young child nutrition (MIYCN) and mobidity   - Standard infant and young child feeding (IYCF) questionnaire and indicators (exclusive breastfeeding, minimum dietary diversity [MDD], minimum meal frequency [MMF], and minimum acceptable diet [MAD] - Minimum dietary diversity for women (MDD‑W) - Child morbidity -diarrhea, cough, and fever | Survey | At all survey waves |
| Shock experience/exposure   - Drought - Locusts - COVID-19 - Flooding - Market shocks - Livelihood disruption - Illness/death etc. - Violence and community conflict, etc. | Survey | At all survey waves |
| Shock preparedness and response   - Various coping strategies - Participation in formal social safety nets and other humanitarian/development activities - Role of informal social capital - Psychosocial well-being, locus of control, and measures of aspiration | Survey | All survey waves |

Completed samples by type of data collection, Samburu County

**Figure S1:** Timing of surveys and seasons.

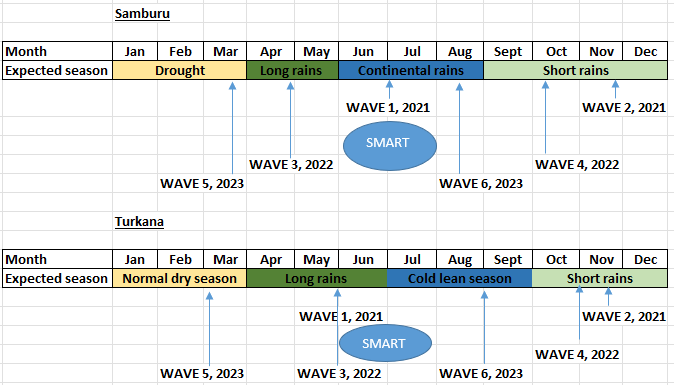
